# Pre-clinical Research of Human Amnion-derived Mesenchymal Stem Cells and its First Clinical Treatment for a Severe Uremic Calciphylaxis Patient

**DOI:** 10.1101/2021.09.23.21261751

**Authors:** Lianju Qin, Jing Zhang, Yujie Xiao, Kang Liu, Yugui Cui, Fangyan Xu, Wenkai Ren, Yanggang Yuan, Chunyan Jiang, Song Ning, Ming Zeng, Guang Yang, Hanyang Qian, Anning Bian, Fan Li, Xiaoxue Ye, Shaowen Tang, Juncheng Dai, Jing Guo, Qiang Wang, Bin Sun, Yifei Ge, Chun Ouyang, Xueqiang Xu, Jing Wang, Yaoyu Huang, Hongqing Cui, Jing Zhou, Meilian Wang, Zhonglan Su, Yan Lu, Di Wu, Zhihong Zhang, Jingping Shi, Wei Liu, Li Dong, Yinbing Pan, Baiqiao Zhao, Ying Cui, Xueyan Gao, Zhanhui Gao, Xiang Ma, Aiqin Chen, Jie Wang, Meng Cao, Qian Cui, Li Chen, Feng Chen, Youjia Yu, Qiang Ji, Zhiwei Zhang, Mufeng Gu, Xiaojun Zhuang, Xiaolin Lv, Hui Wang, Yanyan Pan, Ling Wang, Xianrong Xu, Jing Zhao, Xiuqin Wang, Cuiping Liu, Ningxia Liang, Changying Xing, Jiayin Liu, Ningning Wang

**Author notes:** **Address reprint requests to:** Pro. Ningning Wang at Department of Nephrology, The First Affiliated Hospital of Nanjing Medical University, Jiangsu Province Hospital, Nanjing, Jiangsu, China; or to Pro. Jiayin Liu at State Key Laboratory of Reproductive Medicine, Center of Clinical Reproductive Medicine, The First Affiliated Hospital of Nanjing Medical University, Jiangsu Province Hospital, Nanjing, Jiangsu, China; or to Pro. Lianju Qin at State Key Laboratory of Reproductive Medicine, Center of Clinical Reproductive Medicine, The First Affiliated Hospital of Nanjing Medical University, Jiangsu Province Hospital, Nanjing, Jiangsu, China. Lianju Qin, Jing Zhang and Yujie Xiao contributed equally to this article.

## Abstract

Calciphylaxis is a rare disease characterized histologically by microvessel calcification and microthrombosis, with high mortality and no proven therapy. We reported a severe uremic calciphylaxis patient with progressive skin ischemia, large areas of painful malodorous ulcers and mummified legs. Because of her rapid progression and refractory to conventional therapy, human amnion-derived mesenchymal stem cells (hAMSCs) treatment was approved. Establishment and release inspection of hAMSCs, efficacy and safety assessment including cytokines secretory ability, immunocompetence, tumorigenicity and genetics analysis *in vitro* were introduced. We further performed acute and long-term hAMSC toxity evaluations in C57BL/6 mice/rats, abnormal immune response tests in C57BL/6 mice and tumorigenic tests in the neonatal NU nude mice. After pre-clinical research, she was treated by hAMSCs with intravenous and local intramuscular injection and external supernatants application to her ulcers. When followed up to 15 months, her blood-based markers of bone and mineral metabolism were improved, with regeneration of skin soft tissue and a more favorable profile of peripheral blood mononuclear cells. Skin biopsy after 1 month treatment showed vascular regeneration with mature non-calcified vessels within dermis and 20 months later re-epithelialization restored the integrity of damaged site. No infusion or local treatment related adverse events occurred. To the best of our knowledge, this is the first evidence for the clinical use of hAMSCs. These findings suggest hAMSCs warrant further investigation as a potential regenerative treatment for uremic calciphylaxis with effects of inhibiting vascular calcification, stimulating angiogenesis and myogenesis, anti-inflammatory and immune modulation, multi-differentiation, re-epithelialization and restorage of integrity.

## Introduction

Calciphylaxis is a rare, devastating disorder causing excruciatingly painful ischemic skin lesions due to microvascular calcification and microthrombosis and endothelial injury, which in turn results in infarction of tissues (Nigwekar et al., 2018). The annual incidence among patients with end-stage kidney disease (ESKD) reaches 0.04% (Brandenburg et al., 2017), and sepsis due to the infection of ulcerated wounds is a common cause of death (Seethapathy and Nigwekar, 2019). There is presently no approved therapy related to its rare incidence and poorly understood pathogenesis. In calciphylaxis patients receiving dialysis, 1-year mortality may reach 80% (McCarthy et al., 2016).

Stem cells are reported to be involved in angiogenesis, myogenesis, activation, proliferation, re-epithelialization of wound healing and play an important role in immune modulation, tissue remodeling, and extracellular matrix deposition (Hassanshahi et al., 2019). Human amnion-derived mesenchymal stem cells (hAMSCs) are abundant, greater cell yields at harvest, the presence of a more “youthful” phenotype, compatibility for use in allogeneic transplants, and enhance immunomodulatory properties as compared with human adipose-derived stem cells (hADSCs) and bone marrow-derived stem cells (hBMSCs)(Topoluk et al., 2017). Importantly, hAMSCs have enhanced wound healing properties through differentiation and stimulation of neoangiogenesis (Ertl et al., 2018).

Here we report female uremic calciphylaxis patient in her thirties with large areas of painful malodorous ulcers and legs with a mummified appearance. She had been on peritoneal dialysis (PD) for 5 years and was admitted in our hospital, with multiple skin lesions accompanied by pain for more than 1 month. She had been switched to hemodialysis (HD) because of peritonitis. She had progressive skin lesions, with induration, plaques, purpura, reticularis and ecchymosis on her back, thighs, lower limbs and buttocks. The multiple, bilateral painful malodorous necrotic ulcerations were surrounded with leather-like skin.

Her multiple medical problems included chronic kidney disease (CKD)G5D (defined as estimated glomerular filtration rate (eGFR) of <15 ml/min/1.73m^2^ or requiring dialysis—often called ESKD), calciphylaxis, skin and soft tissue infections, malnutrition, secondary hyperparathyroidism (SHPT), PD-related tunnel infection and hypertension. Treatments included nutritional support, antibiotic administration and infection control, low calcium hemodialysis and continuous renal replacement therapy (CRRT), management of anemia and CKD-mineral and bone disorder (CKD-MBD), sodium thiosulfate intravenously, wound care, pain management, anticoagulation, thrombosis prevention, cardioprotective and gastrointestinal protective therapy, as well as therapy for neurotrophic issues (Seethapathy and Nigwekar, 2019).

Because of rapid progression of the calciphylaxis which was refractory to conventional therapy, written informed consent was provided by the partient prior to receive hAMSC treatment as approved by the ethics committee of The First Affiliated Hospital of Nanjing Medical University, Jiangsu Province Hospital (2018-QT-001). For death of the patient, a statement confirming consent for the publication of her information was provided by legally authorized representatives.

To the best of our knowledge, this is the first evidence for the clinical use of hAMSCs and treatment for a severe uremic calciphylaxis patient. Her complete remission of calciphylaxis suggested further exploration of hAMSC therapy may be indicated.

## Results

### Pre-clinical research of hAMSCs

Quality control and pre-clinical research indexes of hAMSCs are listed in Table1.

**Table 1.**
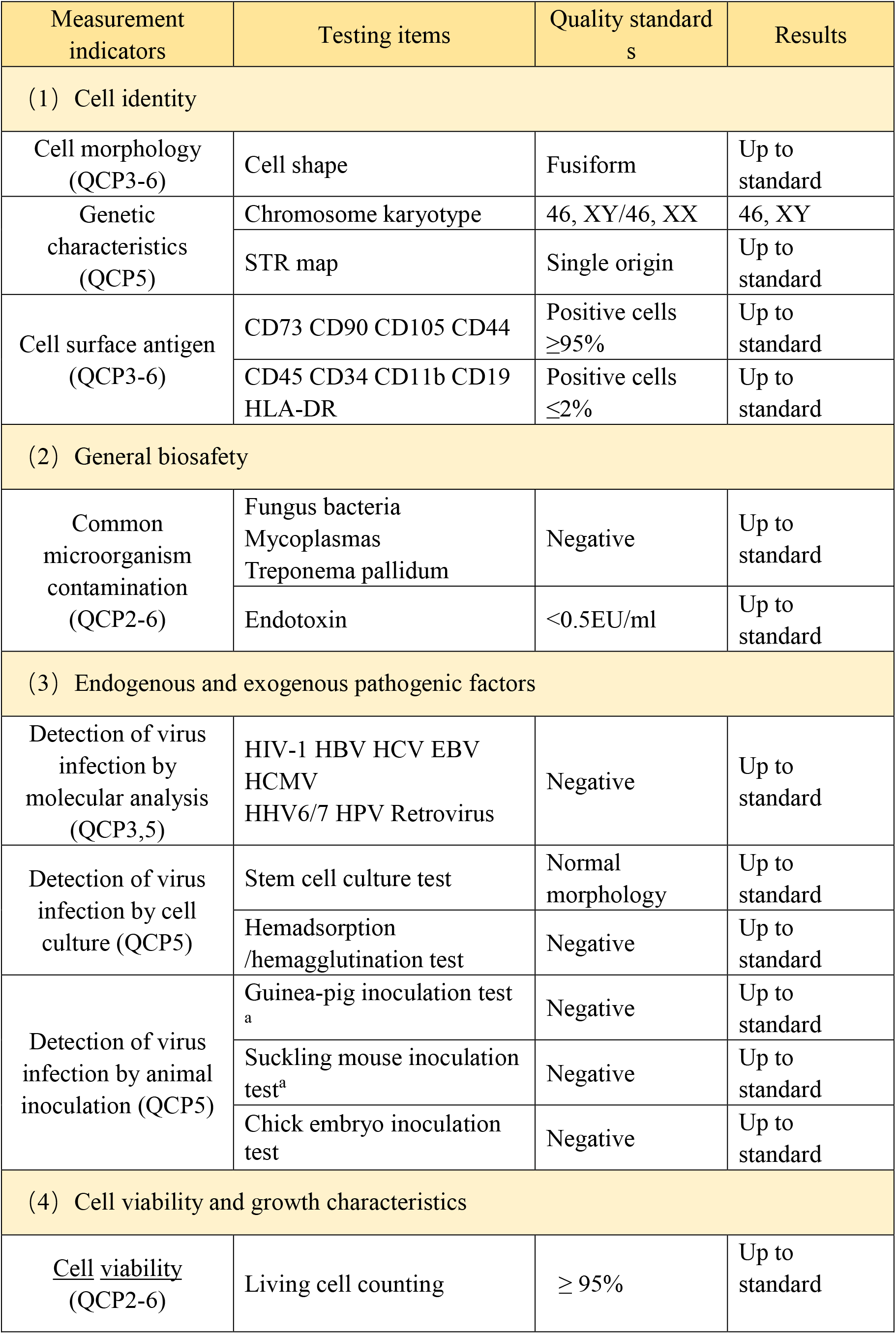
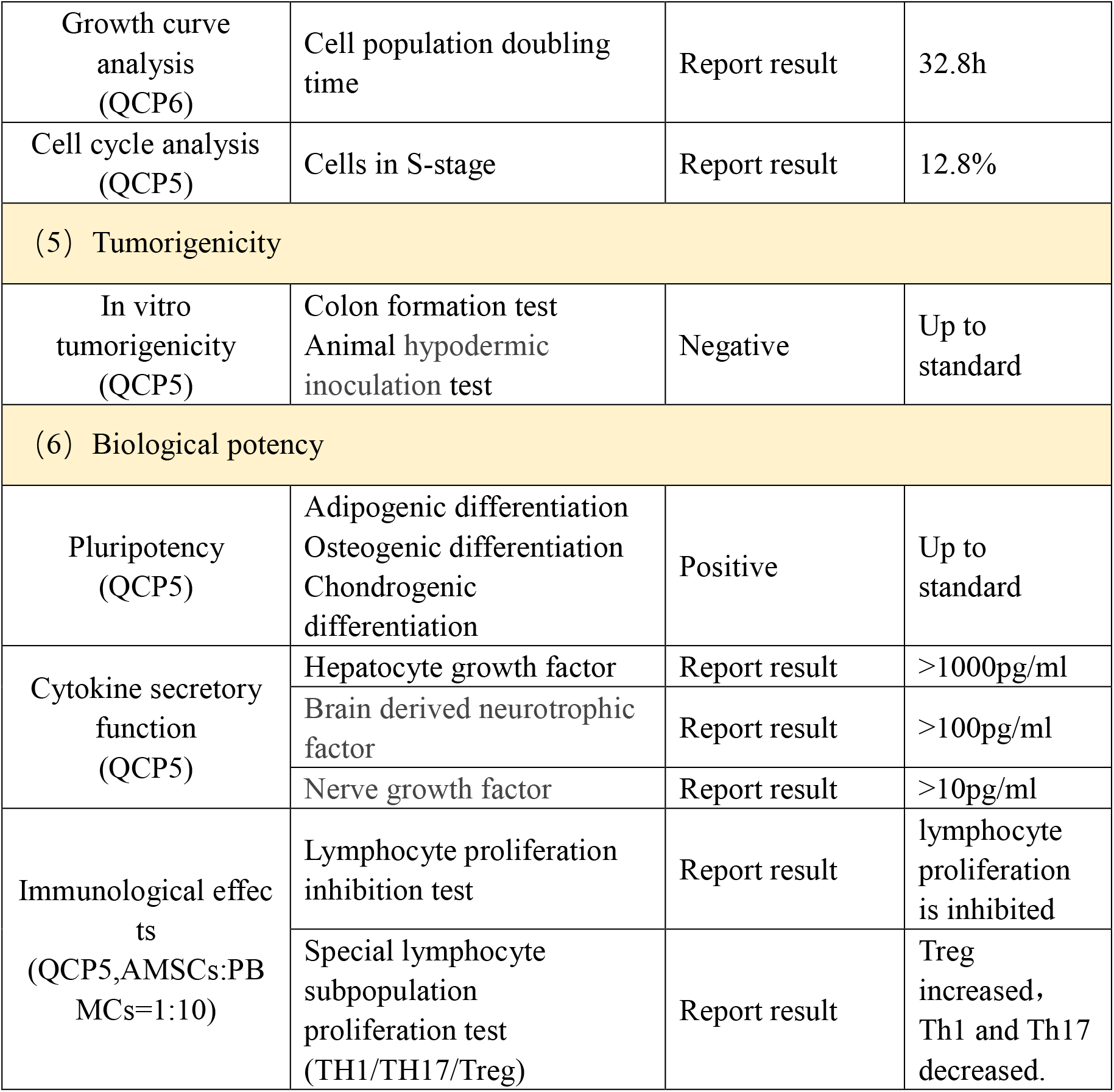
Quality control and pre-clinical research of cell line HAMSC10.

### Pre-clinical release inspections of hAMSCs

Release inspections consisted of general biosafety, viability, growth characteristics were up to standard (Fig 1A). Cells surface antigens including CD44, CD73, CD90 and CD105 positive cells were ≥95%, while CD11b, CD19, CD34, CD45 and HLA-II positive cells were ≤2%, and they were up to quality standard (Fig 1B).

**Figure 1.**
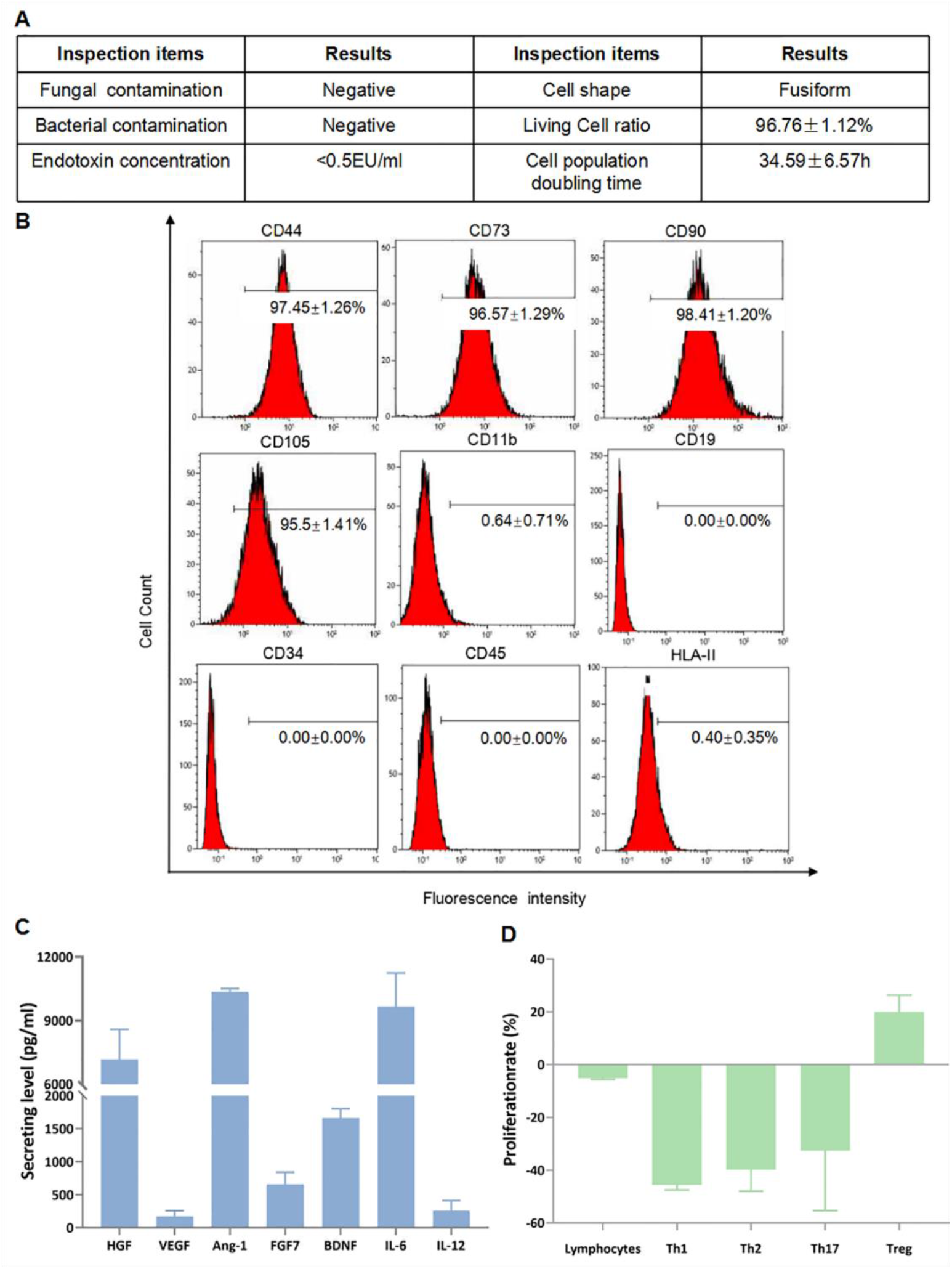
Pre-clinical release inspection and effective assessment. (A) General biosafety and cell viability of hAMSC preparation, they were up to quality standard. (B) Release inspection on hAMSC surface antigens. (C) Analysis for cytokine secretions from hAMSC culture supernatants. (D) HAMSCs showed immunocompetence *in vitro* when co-cultured with lymphocytes from healthy persons.

### HAMSC effective potency of cytokines secretory and immunocompetence function

HAMSC supernatant contains high levels of hepatocyte growth factor (HGF, 7166.2±1425.6 pg/ml), angiogenic factors including angiopoietin-1 (Ang-1, 10337.5±172.3 pg/ml), brain-derived neurotrophic factor (BDNF, 1658.8±144.9 pg/ml), interleukin-6 (IL-6, 9655.5±1588.3 pg/ml), and moderate levels of vascular endothelial growth factor (VEGF, 167.6±89.2 pg/ml), fibroblast growth factors-7 (FGF-7, 654.7±188.2 pg/ml) and interleukin-12 (IL-12, 256.4±158.7 pg/ml)(Fig 1C). The hAMSCs have regulatory functions in immune cells especially T cells, with can slightly inhibit the proliferation rate of total lymphocytes (−5.2±0.4%), strongly inhibit the proliferation of Th1 (−45.5±2.0%), Th2 (−39.8±8.1%) and Th17 cells (−32.6±22.7%), and promote the proliferation rate of Treg cells (20.0±6.3%) (Fig 1D).

### HAMSC safety assessment of tumorigenicity *in vitro* and genetics analysis

The soft agar clone formation experiment was observed for 3 weeks, indicating that the negative control (MRC-5) and hAMSC group had no cloning, while the positive control group (Hela) had obvious colony formation, with the clone number of 232.7±20.4, and the clone formation rate of 7.76±0.68%. It was preliminarily shown that hAMSCs were not tumorigenic *in vitro* (Fig 2A-2F). G-banding analysis showed that the karyotype was normal (46, XY). Short tandem repeat (STR) analysis proved the cells were a single source population (Fig 2G).

**Figure 2.**
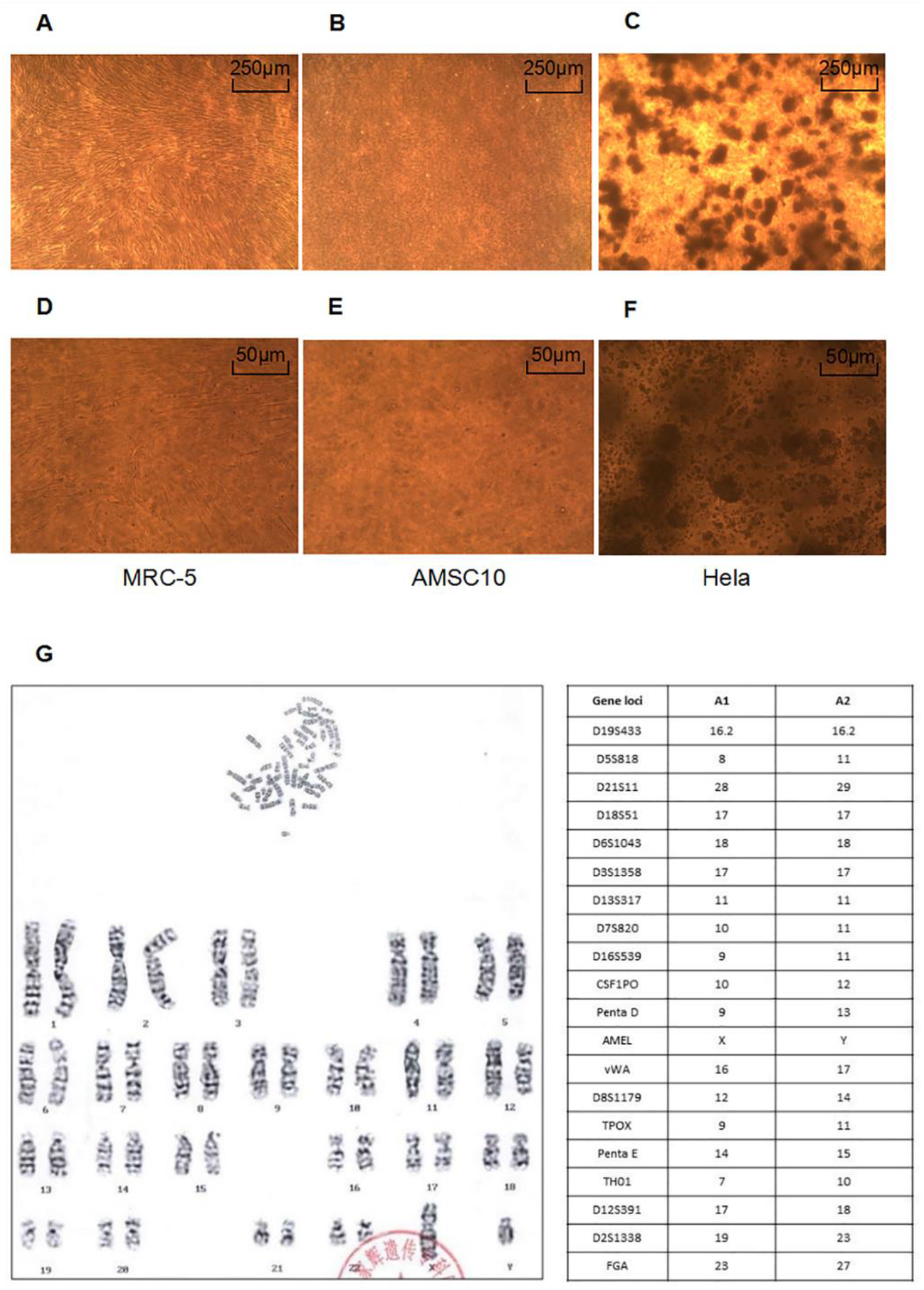
Pre-clinical safety assessment of hAMSCs. The soft agar clone experiment to detect the tumorigenicity of hAMSCs which were cultured for 21 days in *vitro* (A**-**C, ×40; D**-**F, ×200): (A, D) Human normal embryo lung fibroblasts (MRC-5, 3.0×10^3^ cells/well) were negative controls; (B, E) HAMSCs (6.0×10^3^ cells/well) showed no tumorigenic effect; (C, F) Human cervical cancer cells (Hela, 3.0×10^3^ cells/well) were used as positive controls. (G)The karyotype and short tandem repeat (STR) of hAMSC10.

### Maximum tolerated dosages in mice/rats and their estimated equivalent dosages in human

#### Determinations of mice/rats equivalent dosages in human

The dosages conversion between mice/rats and human is not only based on the body mass as biochemical functional systems vary among different species (A. B. Nair and Jacob, 2016). For a man whose body weight is 60 kg and body surface area is 1.53m^2^, his intravenous administration dosage for hAMSCs is 1.0×10^6^cells/kg. Here we used the Mech formula to determine surface area, which is a classical approach to estimate equivalent dosages between human and mice/rats (A. Nair et al., 2018). This formula is expressed as: S = KW^2/3^, in which S is the surface area in square centimeters, W is the weight in kilograms, and K is a constant (K, 1879).

#### Acute toxicity tests of single hAMSC intravenous administration in mice and rats

The maximum tolerated dose in C57BL/6 mice was 7.50×10^7^ cells/kg (approximately 6 times for the clinical intravenous administration dosage, equivalent to 6.0×10^6^ cells/kg in human). High doses of hAMSCs can cause pulmonary vascular embolism and death in mice. The maximum tolerated dose in SD rats was 1.20×10^8^ cells/kg (approximately 20 times for the clinical intravenous administration dosage, equivalent to 20.0×10^6^ cells/kg in human) (Fig 3A).

**Figure 3.**
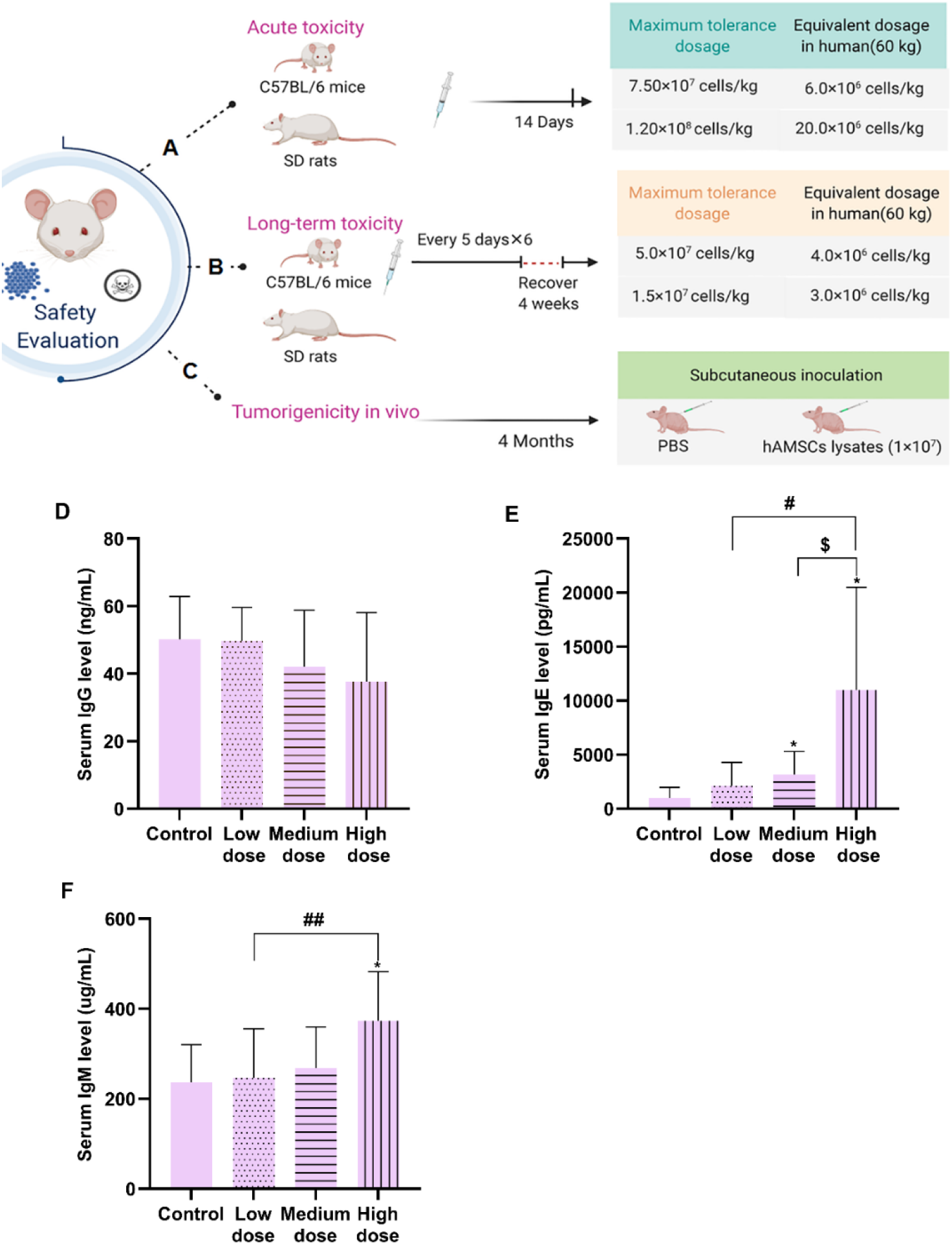
HAMSCs animal tests for maximum tolerated dosage, tumorigenic and abnormal immune response. Maximum tolerated hAMSCs dosage in C57BL/6 mice/rats were investigated with acute toxicity (A) and long toxicity (B) tests, their estimated equivalent dosages in human were also displayed. Tumorigenic tests in newborn nude mice suggested that hAMSCs were not tumorigenic *in vivo* (C). Serum IgG (D), IgM (E) and IgE (F) levels were measured in C57BL/6 mice with abnormal immune response tests.

#### Long term toxicity tests of multiple hAMSC intravenous administration in mice and rats

There was no significant toxicity in C57BL/6 mice with hAMSCs dose of 5.0×10^7^cells/kg (equivalent to 4.0×10^6^ cells/kg in human). No significant toxicity was observed in SD rats with hAMSCs dose of 1.5×10^7^cells/kg (equivalent to 3.0×10^6^cells/kg in human) (Fig 3B).

### Tumorigenic tests of hAMSCs in mice

After 4 months, no tumor or nodules were observed in the neonatal NU nude mice subcutaneously inoculated with hAMSCs lysate at the scapula, suggesting that hAMSCs were not tumorigenic *in vivo* (Fig 3C).

### Abnormal immune response tests in mice with hAMSC intravenous administration

After the first hAMSC intravenous administration, one male mouse in the high-dose group (1.0×10^8^ cells/kg) was died because of pulmonary embolism. The appearance and behavior of surviving mice in each subgroup were normal.

Among control, low-dose, medium-dose and high-dose hAMSC groups, serum IgG levels displayed no significant difference (Fig 3D). Serum IgE levels increased in dose-dependent manner, especially in the medium-dose and high-dose subgroups (Fig 3E). Serum IgM levels showed dose-dependent increasing trend after hAMSC intravenous administration, especially in the high-dose group (Fig 3F).

### Treatment course and dosage of hAMSCs for the uremic calciphylaxis patient

Prior to clinical hAMSC treatment, the patient’s hAMSCs-lysate skin test showed negative. Later, hAMSCs were administered intravenously to our patient at a dose of 1.0×10^6^ cells per kilogram of body weight, local intramuscular injection along the wound edge (2.0×10^4^ cells/cm^2^) and external application of the cell culture supernatant on wound surfaces. The patient’s hAMSC treatment course is outlined in Fig. 4A.

**Figure 4.**
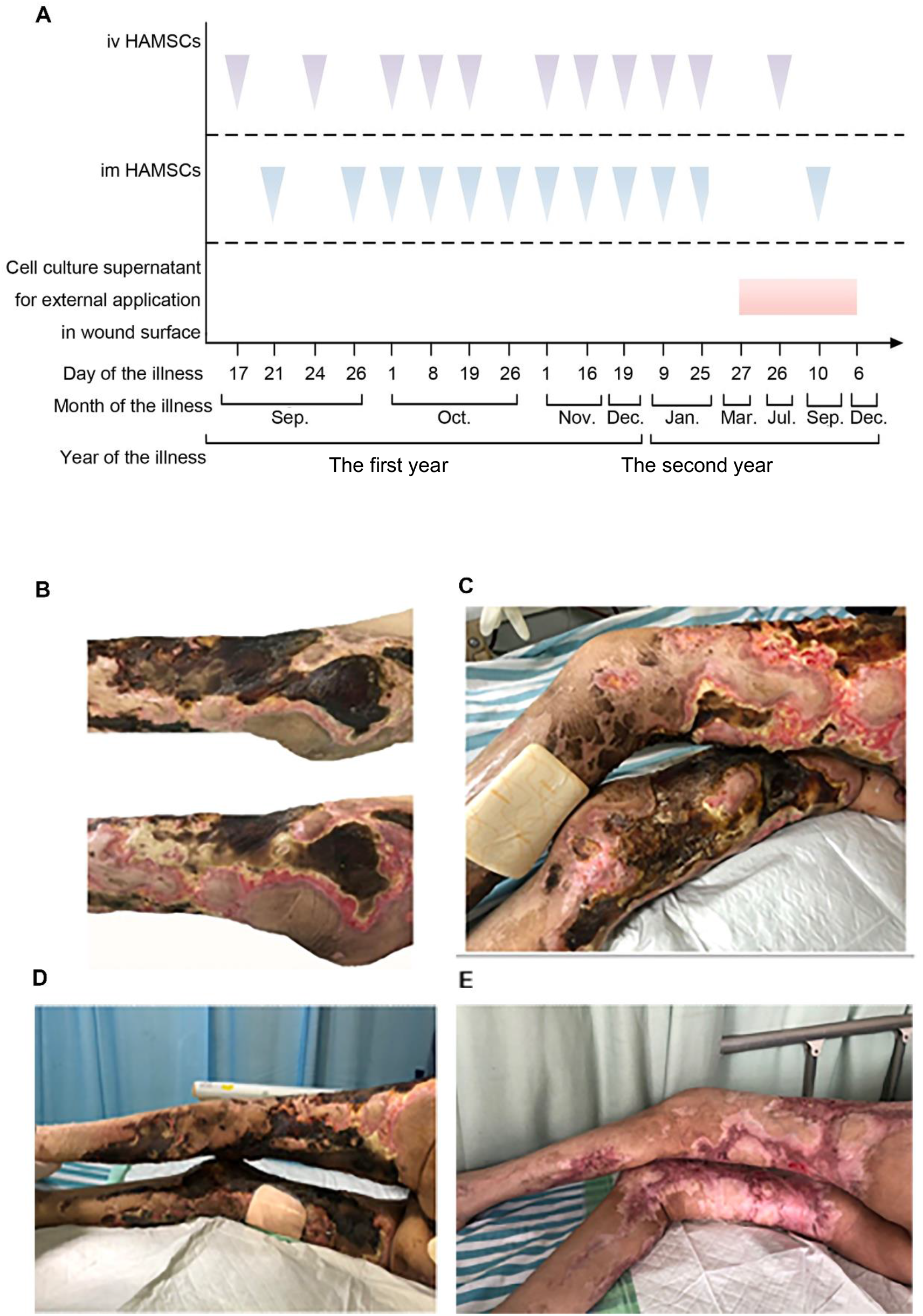
Clinical hAMSC treatment for the uremic calciphylaxis patient. An overview of hAMSC therapeutic schedule for a uremic calciphylaxis patient. Wounds on the lower limbs before hAMSC treatment. After 14 days of intravenous infusion plus local intramuscular injection, the surface of wounds began to improve. (C) The left thigh with local intramuscular hAMSC injection for one month recovered better than the right thigh which without local intramuscular injection. Panel D displayed the wounds of lower limbs before treatment and panel E indicated that the skin lesions were healed when treated with hAMSCs for one year.

### Healing progress of skin lesions treated with hAMSCs

Before treatment, the wounds showed multi-necrotizing ulcers with eschar and infection. After intravenous plus local intramuscular hAMSCs injection for 14 days, the skin lesions had substantially improved (Fig. 4B). During the early stages of calciphylaxis, the patient was in great pain, not fully controlled by analgesics and evident difficulty when it was necessary to change body position. Hence, we first gave intramuscular hAMSC injection only in the left thigh. Then, after one month, when reduced eschar and regenerative tissue were observed, it was clear that the left thigh was healed as compared to the right thigh (Fig. 4C), indicating that local injection could accelerate the wound recovery. Compared with pre-treatment of hAMSCs (Fig. 4D), the lower limbs were healed well after one year (Fig. 4E), and her general condition were improved after 15 months.

### Adverse events

Not infusion or local treatment related adverse events occurred.

### Measurements of blood parameters

After hAMSC treatment, hemoglobin (Hb) and serum albumin levels gradually increased to normal. The levels of leukocytes, platelets, and high-sensitivity C-reactive protein (hs-CRP) decreased substantially, suggesting improvement of inflammation status. A moderate rebound in serum phosphorus and alkaline phosphatase (ALP) levels were observed because of her poor controlled diet. However, serum calcium and iPTH levels remain in normal treatment target (Fig. 5A-5I). Furthermore, serum tumor markers were stable in the normal range after hAMSC treatment for 15 months (Fig. 5J-5M).

**Figure 5.**
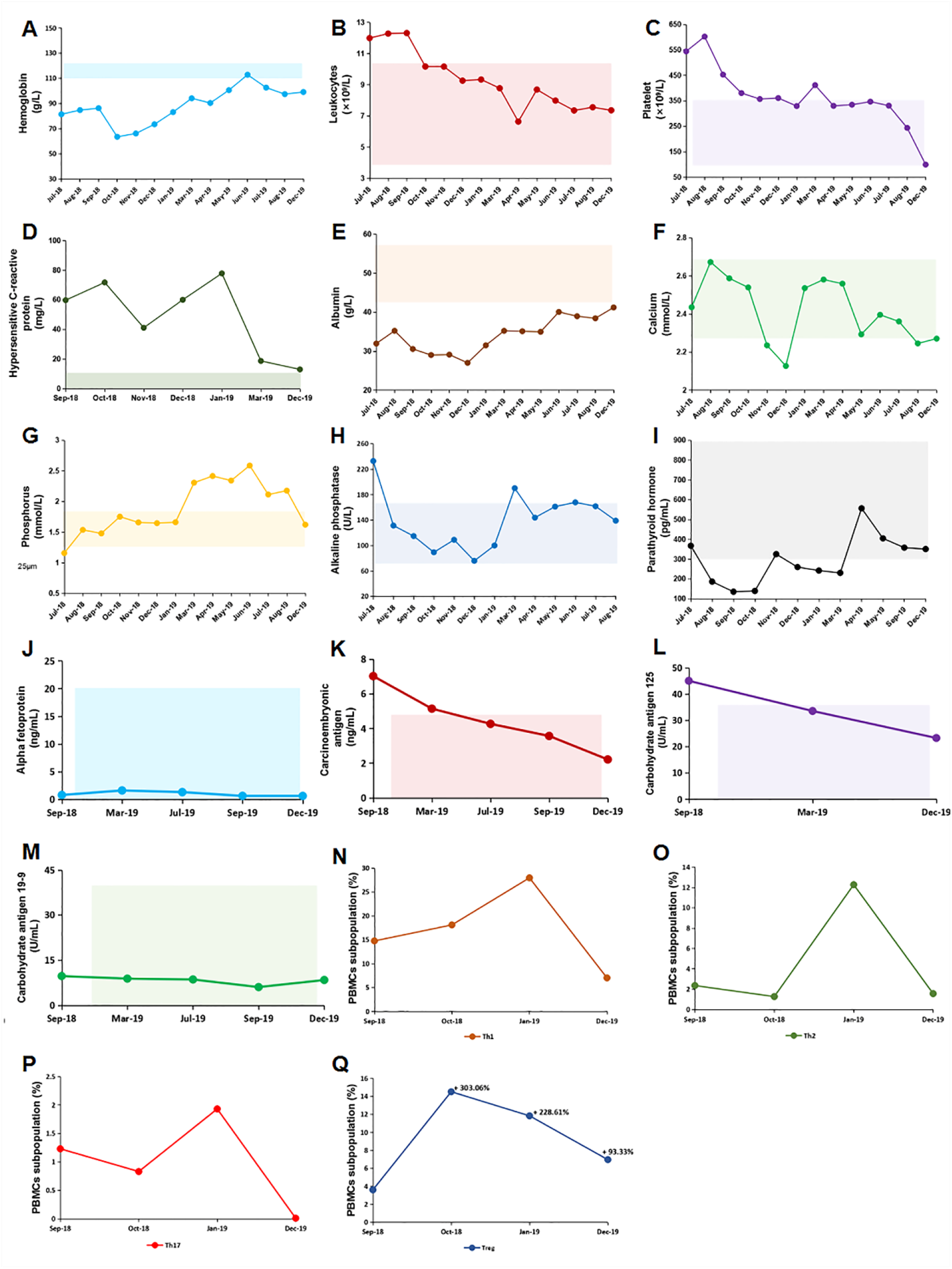
The laboratory data of uremic calciphylaxis patient before and after hAMSC treatment. After hAMSC treatment, we tested (A) Hemoglobin, (B) leukocytes, (C) platelets, (D) hypersensitive C-reactive protein, (E) serum albumin, (F) serum calcium, (G)serum phosphate, (H)serum alkaline phosphatase, (I) serum parathyroid hormone, (J)serum alpha fetoprotein, (K)serum carcinoembryonic antigen, (L) serum carbohydrate antigen 125, (M) serum carbohydrate antigen 19-9. Shaded areas showed reference ranges. PBMCs subpopulation of patient during the course of hAMSC treatment including Th1 (N), Th2 (O), Th17 (P) and Treg (Q) were detected.

### Subpopulations of peripheral blood mononuclear cells (PBMCs) after hAMSCs treatment

The patient had elevated baseline Th1/Th2/Th17 levels, higher Th1/Th2 ratio (6.25), and lower Treg level. After the first month of hAMSC treatment, Th1 and Treg cells increased (22.71% and 303.06%, respectively), while Th2 and Th17 cells decreased (−45.76% and -32.52%, respectively). After 15 months of hAMSC treatment, Th1/Th2/Th17 cells were repressed (−52.41%, -33.47% and -99.19%, respectively) and the proliferation of Treg cells were enhanced (93.33%). Importantly, she had improved Th1/Th2 ratio (4.47:1), which was closer to the normal value (2:1) after hAMSC treatment for 15 months (Fig. 5N-5Q).

### Histological characteristics of skin biopsy before and after hAMSC treatment

The skin biopsy before hAMSC treatment revealed calcification, fibrointimal hyperplasia of dermal and subcutaneous arterioles, leading to thrombosis in microvessels of the subcutaneous adipose tissue and dermis, accompanied by cutaneous ischemia and necrosis. The calcified deposits are pure calcium apatite, located within the media and intima of blood vessels, and were also found in the adipocytes and collagen fibers (Fig. 6A-6B). After treatment, there was evidence of vascular regeneration, with mature non-calcified blood vessels identified within the dermis (Fig. 6C). This process, termed re-epithelialization, enables the generation of a new epidermis, which restores the integrity of the damaged site (Fig. 6D).

**Figure 6.**
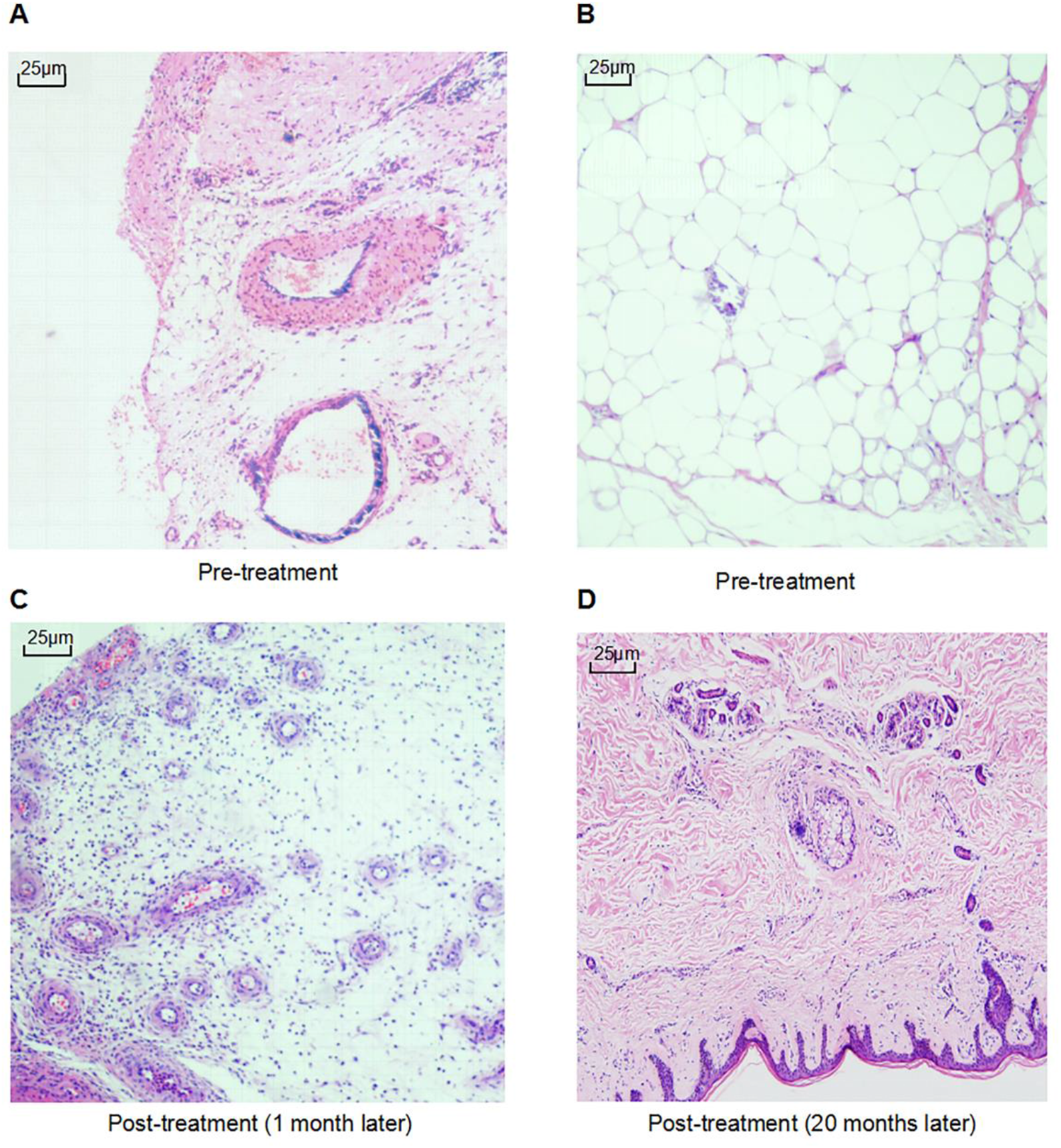
Skin pathology analysis during the course of hAMSC treatment in the uremic calciphylaxis patient. (A) Calcification of the middle layer of artery, necrosis and inflammation before hAMSC treatment. (B) Calcification of adipose tissue before hAMSC treatment. (C) Regenerated mature blood vessels after hAMSC treatment for 1 month. (D) The regenerated skin tissues restored the integrity of lesion site after hAMSC treatment for 20 months (Hematoxylin-Eosin staining, ×400).

## Discussion

This uremic patient was diagnosed as calciphylaxis based on typical clinical manifestations with skin lesions, severe pain and skin histology. She was refractory to traditional therapies including intravenous sodium thiosulfate (Peng et al., 2018), wound care, pain management, anticoagulation, thrombosis prevention, the co-interventions of lowered dialysate calcium and increased length or frequency of dialysis sessions, etc (Nigwekar et al., 2018).

Arteriolar calcification due to VSMCs phenotype transdifferentiation plays a crucial role in the pathogenesis of calciphylaxis (Nigwekar et al., 2018). Intravenouse adipose-derived MSCs (ASCs) treatment has been proved to suppress thoracic arterial medial calcification in CKD rats (Yokote et al., 2017). BMSC-derived exosomes could inhibit high phosphate-induced aortic calcification by decreasing high mobility group box 1 (HMGB1) level via the Sirtuin 6-HMGB1 deacetylation pathway (Wei et al., 2021). BMSC-derived exosomes inhibited high phosphorus-induced VSMCs calcification *in vitro* by modifying miRNA profiles involved in the Wnt, mTOR, or MAPK signaling pathway (Guo et al., 2019)and regulating NF-κB axis (Y. Liu et al., 2021a).

Except for beneficial to the treatment of vascular calcification, stem cells can promote neovascularization by multi-differentiation into keratinocytes, endothelial cells, vascular smooth muscle cells, and act as perivascular cells. Paracrine secretions of cytokines, growth factors, microvesicles/exosomes and chemokines can modulate angiogenesis, apoptosis, immune responses and facilitate the regeneration of damaged tissues (L. Zhao et al., 2017; A et al., 2019).

Here we proved hAMSC supernatant contained high levels of cytokines. As an upstream-indirect proangiogenic growth factor, HGF stimulates endothelial cells to proliferate and migrate while inhibiting cells apoptosis and tissue reconstruction (Pang et al., 2018), inducing VEGF and blood vessel formation (D. Jiao et al., 2016a). HGF is also a potent immunomodulatory and anti-inflammatory factor (Maraldi et al., 2015; Pang et al., 2018). Ang-1 is reported to accumulate in the extracellular matrix, enhancing recruitment of pericytes and smooth muscle cells, promoting endothelial cells survival, proliferation, migration and angiogenesis (Bupathi et al., 2014).

BDNF stimulates new vessels formation by increasing VEGF (Halade et al., 2013), regulates the primary pro-inflammatory transcription factors such as NF-κB, and restricts the magnitude of inflammatory response (Qin et al., 2019). Muscle-derived BDNF plays a role in muscle repair, regeneration, and differentiation (Pedersen, 2013). IL-6 has mitogenic and proliferative effects on keratinocytes and is chemoattractive to neutrophils (Barrientos et al., 2008; Cañedo-Dorantes and Cañedo-Ayala, 2019). As a pro-angiogenic factor, IL-6 can regulate endothelial progenitor cells migration (Fan et al., 2008), activate quiescent microvascular endothelial cells and induce the formation of tubular structures (Potente et al., 2011; Wang et al., 2012). IL-6 is essential for maturation, proliferation, differentiation, and maintenance of B cells/plasma cells and proinflammatory Th17 and Th2 cells (Tvedt et al., 2017). Furthermore, IL-6 is linked with muscle stem cells and have myogenesis effects (Pedersen, 2013).

VEGF affects most downstream activities of angiogenesis including endothelial cells proliferation, migration, invasion, survival and promotes vascular permeability (Waldner et al., 2010; Hu et al., 2019). VEGF also induces extravascular leakage of plasma proteins, modulates extracellular matrix proteolysis (Bien et al., 2015), regulates the local immune response and helps recovery from inflammation (Cañedo-Dorantes and Cañedo-Ayala, 2019). FGF-7 can contribute to synthesize various pro-angiogenic molecules, including VEGF, to promote angiogenesis (Qu et al., 2018). As keratinocyte growth factor, FGF-7 is upregulated after skin injury, stimulates the proliferation (Kao et al., 2011) and migration (Qu et al., 2018) of keratinocytes and accelerates wound healing by reepithelialization (Iwamoto et al., 2015). FGF7 is important for the formation of hair follicles (Qu et al., 2018). IL-12 induces Th0 to Th1 cells to enhance the cellular immunity (Zhou et al., 2019), promoting the cytotoxic activity of NK and T lymphocytes, increasing the secretions of cytokines including interferon-gamma (IFN-γ) and tumor necrosis factor alpha (TNF-α)(J. Zhao et al., 2020).

MSCs can regulate both innate and adaptive immunity by modulating activation, maturation, proliferation or cytolytic activity of multiple immune cells, including T, B and natural killer lymphocytes, dendritic cells, monocytes and macrophages (Y. Shi et al., 2018). In this calciphylaxis patient, an increase in the Th1/Th2 ratio at baseline indicated that inflammation reactions were triggered and Th1/Th2 balance was broken (Chi et al., 2019). After 15 months of hAMSC treatment, her reduced Th1/Th2 ratio revealed the suppression of inflammation and improved health. The anti-inflammatory properties of MSCs have been linked to their immunoregulation potential (Mishra et al., 2020). Consistent with previous studies (Y. Shi et al., 2018; Harrell et al., 2020), we proved that hAMSCs can suppress the proliferation and differentiation of Th1/Th17 cells, induce functional Treg cells and promote cell regeneration.

The use of hAMSCs in animal models with various diseases have been documented previously. Long-term effects of intravenous hAMSC administration for amyotrophic lateral sclerosis were investigated in SOD1^G93A^ mice, after treated with hAMSCs (1×10^6^ cells) at 12, 14 and 16 weeks, the mice displayed retarded disease progression and extended survival (Sun et al., 2014). HAMSC (1×10^6^ cells) administrated intravenously can ameliorate spatial learning and memory function in C57BL/6J-APP transgenic mice with Alzheimer’s disease after 3-week treatment (H. Jiao et al., 2016b).

HAMSCs are also reported to treat skin lesions with local injection. C57BL/6 mice with deep second-degree burn on skin were injected with hAMSCs subcutaneously (2×10^6^) near the wound, hAMSCs and conditional medium can cure skin injury by inhibiting apoptosis and promoting their proliferation (Li et al., 2019). In C57BL/6 mice with full-thickness skin wound, hAMSCs subcutaneously injected(1×10^6^) along the wound edge can contribute the macrophages transformation from M1 to M2 type, induce up-regulation of anti-inflammatory and anti-fibrotic factors, down-regulate inflammation mediated factors (C. S. Shi et al., 2020). In SD rats with full-thickness skin defect wound, hAMSCs injected at 0.5 × 10^4^ cells, 0.5 × 10^5^ cells, and 0.5 × 10^6^ cells were proved to promote wound healing and epithelialization after 15 days (Gao et al., 2020). In SD rats defecting in the middle of tibialis anterior muscle, gelatin methacryloyl hydroge with hAMSCs (5×10^5^/mL) was implanted into the defect area, which displayed agglomeration of cells after 2 weeks (Zhang et al., 2019).

Based on above references about intravenous/local hAMSC injection and our animal experiments for maximum tolerated dosages, we designed hAMSC therapeutic schedule for this uremic calciphylaxis patient with the approval of our hospital ethics committee. The patient was treated with hAMSCs by intravenous and local intramuscular injection within safe dosage. Furthermore, hAMSCs’ supernatant was applied on her ulcers. She can tolerate well with skin regeneration. Skin biopsy from her thigh showed viable skin with revascularization without calcification after hAMSC treatment for 1 month and integrated skin tissues after hAMSC treatment for 20 months. However, the patient died due to cerebral hemorrhage in May, 2020. We speculate the death wasn’t directly related to stem cells treatment because intravenous and intramuscular injections had been stopped for 8 months at the time of her death.

In conclusion, hAMSC treatment for this patient is proved to promote skin and soft tissue repair, which are speculated to rely on inhibiting vascular calcification, stimulating neovascularization and myogenesis, anti-inflammatory and immunoregulation potential, multi-differentiation, re-epithelialization and restorage of integrity. This case provides evidence and rationale for the use of hAMSCs as a novel candidate for regenerative treatment in uremic calciphylaxis patients.

## Materials and Methods

### Preparation and quality control of produced hAMSCs

HAMSCs were prepared in State Key Laboratory of Reproductive Medicine, The First Affiliated Hospital of Nanjing Medical University (H. Liu et al., 2021b), a Good Manufacturing Practice (GMP) compliant laboratory according to the national principle (“Guiding Principle for Cell Therapy Products Research and Assessment Technique,” 2017).

Human amniotic membranes were donated by healthy pregnant women in their 20s or 30s who were undergoing C-sections for full term pregnancies and who provided written informed consent (Fig 7, QCP1), which were given an ethical approval by the ethics committee of The First Affiliated Hospital of Nanjing Medical University, Jiangsu Province Hospital (2012-SR-128). Amniotic membranes that screened negative for microbial contamination were cultured (Fig 7, QCP2). Primary hAMSCs (P0) were cryopreserved as primary cells in our cell bank if negative for microbial and virus contamination (Fig 7, QCP3). P0 cells were passaged three times, cryopreserved as secondary cells bank after confirmed negative microbial contamination (Fig 7, QCP4). Safety and efficacy assessments (Fig 7, QCP5) included microbiology, virology, viability, cells surface antigen, morphology, tumorigenicity, secretory ability and immunocompetence analysis, genetics analysis of karyotype and short tandem repeat (“Guiding Principle for Stem Cell-based Medicinal Products Quality Control and Pre-clinical Research,” 2015; “Guiding Principle for Cell Therapy Products Research and Assessment Technique,” 2017). The hAMSCs passed the quality verification from National Institutes for Food and Drug Control of China (Reports of SH201905141 and SH201905141). Secondary cells bank produced fresh hAMSCs after going through the release inspection (Fig 7, QCP6). Establishment and quality control of hAMSC lines were introduced below (Alviano et al., 2007; “Guiding Principle for Stem Cell-based Medicinal Products Quality Control and Pre-clinical Research,” 2015).

**Figure 7.**
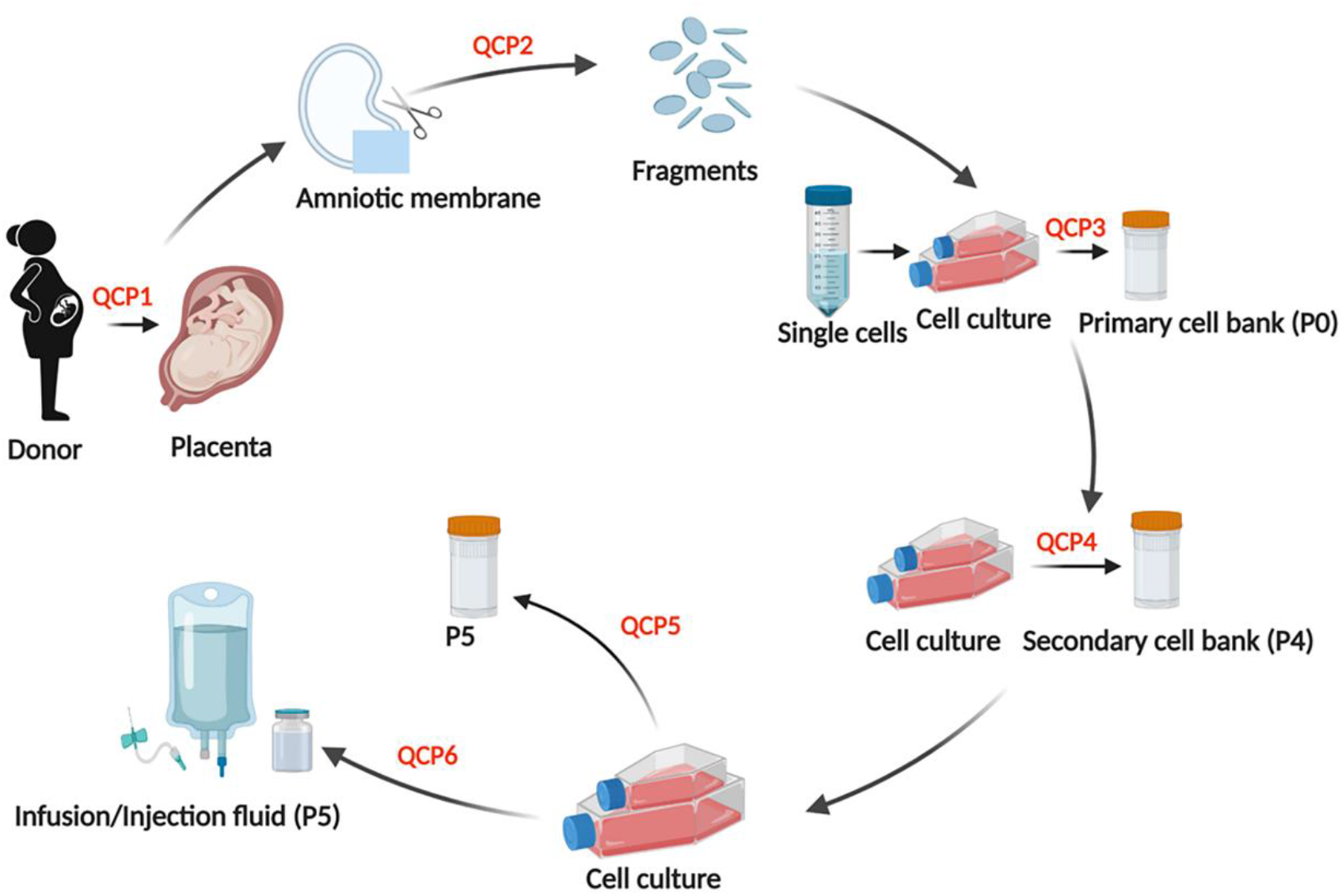
The flowchart of manufacture and pre-clinical quality control for hAMSCs. Quality control point 1 (QCP1), at which virus infection (HIV-1, HBV, HCV, EBV, HCMV, HHV6/7, HPV and retrovirus) were tested, medical history was inquired (excluded donors with contraindication), fetus healthy and body weight were assessed. QCP2, at which microbial contaminations (fungus, bacteria, mycoplasma and treponema pallidum) were tested. QCP3 and QCP4, at which microbial contamination and levels of surface markers were analyzed. QCP5, at which karyotype and STR were confirmed, microbial contamination, cell viability and levels of surface markers were tested, meanwhile oncogenicity *in vitro*, cytokines secretory ability and immunocompetence effects of hAMSCs were analyzed. QCP6, at which microbial contamination, cell shape, cell viability, cell growth and levels of surface markers were analyzed.

### Establishment of hAMSC Lines

Human amniotic membranes were digested by CTS™ TrypLE™ Select Enzyme (GIBCO, Mass, USA) and Collagenase NB 6 GMP Grade (Serva GmbH, Heidelberg, Germany). Digested cells were plated in 15cm dishes and cultured in a 37°C, 5% CO_2_ incubator for 4-5 days. The complete culture medium (CCM) was composed of 5% UltraGRO-hPL (Helios Bioscience, Dublin, Ireland), MEM-alpha (GIBCO, Mass, USA), 1% L-Glutamine (GIBCO, Mass, USA) and 2U/ml heparin sodium. Cells that reached 80-90% confluence were digested by CTS™ TrypLE™ Select Enzyme. The harvested cells were cryopreserved or passaged further to perform the quality test.

### Release inspections on hAMSCs

HAMSCs specific surface markers were detected by flow cytometry. Fungal and bacterial contamination was tested by culture methods according to the national principle (“Guiding Principle for Cell Therapy Products Research and Assessment Technique,” 2017), and bacterial contamination was further assessed by endotoxin detection kit (Zhanjiang Bokang Marine Biological Co., Ltd, China). Mycoplasma infection was assessed by real-time fluorescence quantitative polymerase chain reaction with specific primers (Forward: GGGAGCAAACAGGATTAGATACCCT; Reverse: TGCACCATCTGTCACTCTGTTAACCTC) on an Applied Biosystem® CO. Ltd. machine (QuantStudio™ 7 Flex Real-Time PCR System). Cell viability was assessed by living cell counting. Cell population doubling time was calculated based on living cell counting and equation of T_D_=*(t* × log2)/(log*N*_*t*_-log *N*_*0*_) (*t*:cell culturing time; *N*_*0*_: cell number at the time of planting; *N*_*t*_: cell number after *t* hours growing).

### Analysis for the surface markers of hAMSCs

Cells that reached 80-90% confluence were digested with Tryple (GIBCO, Mass, USA), washed with PBS, and divided into 1×10^5^/10ul/tube. The target antibodies (Table S1) were added, placed at room temperature for 20 minutes and protected from light. The cells were washed with DPBS (GIBCO, Mass, USA), re-suspended, transferred to a flow tube and tested on the machine (Gallios, Beckman-Coulter, France).

### Analysis for cytokines secretions from hAMSC supernatants

When hAMSCs reached 80-90% confluence *in vitro*, the culture supernatants were analyzed by ELISA. Kits from Fcmacs Biotech Co., Ltd. (Nanjing, China) were applied to detect the levels of HGF, BDNF, IL-6 and IFN-γ. VEGF, Ang-1, FGF7 and IL-12 were measured with kits from Mutisciences Biotech Co., Ltd. (Hangzhou, China).

### Analysis of hAMSC immunocompetence *in vitro*

PBMCs were isolated from whole blood of healthy control/patient and co-cultured with hAMSCs at 10:1 for 72 hours, then brefeldin A (BioLegend, CA, USA) was added to stimulate for 5 hours, and cells were collected. Total lymphocytes were labeled with anti-human CD3 (BioLegend, CA, USA) and CD8 antibodies (BioLegend, CA, USA) and Th1/Th2/Th17 cells were labeled with anti-human CD3, CD8, IL-4 (BioLegend, CA, USA), IFN-r (BioLegend, CA, USA) and IL-17A antibodies (BioLegend, CA, USA). Treg cells were labeled with anti-human CD4 (BioLegend, CA, USA), CD25 (BioLegend, CA, USA) and FOXP3 antibodies (BioLegend, CA, USA). All cells were detected by flow cytometry.

### HAMSC tumorigenicity assessed by soft agar experiment *in vitro*

HAMSCs (AMSC10.2.2, P5) were inoculated into soft agar (6-well plates) at low (1.5×10 ^3^/well), medium (3×10 ^3^/well), and high (6×10 ^3^/well) concentrations (n=3 in each group). Human normal embryo lung fibroblasts were negative controls and human cervical cancer cells were positive controls, inoculating into soft agar at 3×10 ^3^/well. Cells were cultured in a 37°C, 5% CO _2_ incubator for 3 weeks, counted the number of clones with more than 20 cells in each group, and calculated the clone formation rate.

### The karyotype and short tandem repeat analysis

Chromosomes were visualized by Giemsa staining. A minimum of 50 metaphases hAMSCs /each sample performed a full karyotype (Catalina et al., 2007; Catalina et al., 2008). Chromosome DNAs were extracted from hAMSCs, multiplex PCR was performed for STR regions of interest(Baine and Hui, 2019). Twenty loci and amelogenin for sex determination were tested.

### Acute toxicity tests of single hAMSC intravenous administration in mice and rats

C57BL/6 mice (6-8 weeks old, n=10 in each group) were injected with hAMSCs via tail vein. A total of 8 doses (be equivalent to 2-32 times of clinical dose with 2-32×10 ^6^ cells/kg in human) were tested. SD rats (6-12 weeks old, n= 4 in each group) were injected with a total of 9 doses (be equivalent to 7-53 times of clinical dose with 7-53×10 ^6^ cells/kg in human). The survival times were observed for 14 days.

### Long term toxicity tests of multiple hAMSC intravenous administration in mice and rats

C57BL/6 mice (7-8 weeks old, n=10 in each group) and SD rats (9-10 weeks old, n=12 in each group) were injected with hAMSCs via tail vein every 5 days for 6 times. They have 4 weeks recovery after the last hAMSC administration.

### Tumorigenic tests of hAMSCs in mice

Tests for tumorigenesis were performed in neonatal NU nude mice aged 3 days by subcutaneously inoculated with hAMSCs lysate (1×10 ^7^ cells /mouse) at the scapula area (n=16), and PBS was injected as negative control(n=11). The mice were observed for 4 months.

### Abnormal immune response tests in mice with hAMSC intravenous administration

C57BL/6 mice (7-8 weeks-old, 16-20g for females and 18-21g for males) were randomly divided into control group (1% human serum albumin), hAMSC low-dose subgroup (2.5×10^7^ cells/kg), hAMSC medium-dose subgroup (5.0×10^7^ cells/kg), and hAMSC high-dose subgroup (1.0×10^8^ cells/kg). There were 10 mice in each group with equal numbers of female and male. The hAMSCs were administered by tail vein, and repeated every 5 days for a total of 3 times. With 4 weeks recovery after last administration, serum IgG, IgM and IgE levels were measured by ELISA (Fcmacs Biotech Co., Ltd., Nanjing, China).

### Measurement of blood parameters for the uremic calciphylaxis patient

Venous whole blood samples were drawn in the morning from the patient after overnight fasting. Routine blood tests were performed using an LH-750 Hematology Analyzer (Beckman Coulter, Fullerton, CA, USA). Biochemical indices were measured using an automatic biochemical analyzer (AU5400; Olympus Corporation, Tokyo, Japan). Serum iPTH levels were measured using a UniCel DxI800 Access Immunoassay System (Beckman Coulter, Fullerton, CA, USA). Serum tumor parameters were measured with the Cobas e602 electrochemiluminescent immunoassay instrument (Roche, Sandhofer Strasse, Germany).

## Data Availability

All the data will be publicly released upon publication.

## Acknowledgements

The authors thank the patient, her family and clinicians who assisted with the data collection. This study was supported by International Society of Nephrology (ISN) Mentorship Program and we thank Professor Marcello Tonelli (University of Calgary, Canada) for his helpful comments and revision on draft of the manuscript.

