## Supplemental Table 1 for "Pre-clinical Research of Human Amnion-derived Mesenchymal Stem Cells and its First Clinical Treatment for a Severe Uremic Calciphylaxis Patient"

**Table S1. Target antibodies for the surface markers of hAMSCs.**

| Antibody Name | Antibody Brand and Item NO. |
|---|---|
| Isotype Control | BD:555573+BioLegend:400314 |
| PE-CD44+FITC-HLA-DR | BD:555479+BD:562008 |
| PE-CD73 | BD:550257 |
| PE-CD90 | BD:555596 |
| PE-CD105 | BD:560839 |
| PE-CD11b | BD:555388 |
| PE-CD19 | BD:555413 |
| PE-CD34 | BD:555822 |
| PE-CD45 | BD:555483 |

PE: Polyethylene; FITC: Fluorescein isothiocyanate; BD: Becton, Dickinson and Company (New Jersey, USA)
